# Early child development in England: cross-sectional analysis of ASQ^®^-3 records from the 2-2^½^-year universal health visiting review using national administrative data (Community Service Dataset, CSDS)

**DOI:** 10.1101/2024.09.28.24314205

**Authors:** Jayu Jung, Sarah Cattan, Claire Powell, Jane Barlow, Mengyun Liu, Amanda Clery, Louise Mb Grath-Lone, Catherine Bunting, Jenny Woodman

## Abstract

**Introduction:** The Ages & Stages Questionnaire (ASQ^®^; 3rd Edition) is a tool to measure developmental delay for children aged between 1 - 66 months which was originally developed in the United States (USA). This measure has been collected in England since 2015 as a part of 2-2^1/2^-year Health Visiting review. However, the quality of the data is known to be incomplete and to-date there have not been any analyses of this data across England looking at disparities between groups of children.

**Objectives:** We identify a subset of child development data, ASQ^®^-3 in Community Service dataset (CSDS) that is sufficiently complete to carry out research then using this dataset we describe child development at aged 2-2^1/2^ years in England (2018/19-2020/21).

**Methods:** We compared counts of ASQ^®^-3 records in CSDS by local authority and financial quarter against the Health Visitor Service Delivery Metrics (HVSDM) to identify a subset of CSDS data that were complete. We described child development using this subset of the data. We used both binary cut-off variable indicating whether a child reached expected/above level of development and continuous ASQ^®^-3 score variable to understand child development at age 2-2^1/2^.

**Results:** Among the 226,505 children from 64 local authorities in our sample, 86.2% met expected level of development. Children from the most deprived neighbourhoods (82.6%), the Black ethnic group (78.9%) and boys (81.7%) were less likely to meet expected level of development. Gender disparity on child development was strong as boys (86.0) in the least deprived neighbourhood were less likely to reach expected level of development compared to the girls (88.2%) from the most deprived neighbourhoods.

**Conclusions:** In order to fully understand child development in England, first ASQ^®^-3 data flow needs to be improved. Second, ASQ^®^-3 data needs to be standardised and validated in the UK context. Developmental support is needed for at least 13.8% of the children who did not meet the expected level of development and especially for those who lived in the most deprived neighbourhoods and boys.

## Introduction

Previous studies have stressed the importance of early child development which in a long term affects educational attainment and is eventually linked to people’s economic, social and health outcomes (1, 2). In line with this evidence, the government has also recognised the importance of reducing early childhood inequalities and releasing services young children(3-6). For example, Healthy Child Programme in England comprises a universal preventative service for children under 5 along with targeted support for families with higher need in order to promote health and wellbeing and reduce inequalities in early childhood (7-10). The Healthy Child Programme is led by health visitors, specialist community public health nurses who lead teams of community staff nurses, nursery nurses, health care assistants, and other specialist health professionals (8, 9, 11).

Health visiting is an important aspect of supporting child development in England which is led by health visiting teams. There are five mandated universal health reviews: during the third trimester of pregnancy, when the child is age 10-14 days (new birth visit), 6-8 weeks (6-8-week review), 12 months (one-year review), and 2-2½ years (2-2½-year review). Each of these mandated contacts has a schedule of health promotion activities and a review of health and development of the child within their family context (12). The 2-2½-year review is the final universal contact between families and the health visiting team before a child starts school at age four: a key part is a review of child development to identify any additional support to be ready for school entry. At this review, a measure of child development called the Ages & Stages Questionnaire (ASQ^®^; 3rd Edition adapted for use in England), is used routinely to collect population level data on early child development for monitoring trends and disparities (13). The ASQ^®^-3 is a tool to measure early child development which was originally developed in the United States (USA) by Squires and colleagues (14) in order to screen developmental delay of young children aged between 1 months and 5 years (10).

Although the ASQ^®^-3 was developed largely based on the norms of the USA, it has also been used in many other countries. In the USA, the ASQ^®^-3 was mainly used for identifying developmental delays especially in medical settings aiming to enhance early detection, clinician-referral, and interventions while it was mainly used for identifying developmental– behavioural differences in intervention/exposure-based cohorts in Scandinavian countries (15). ASQ^®^-3 was also used to screen child development in the European countries (16, 17), countries in Africa (18), South America (19) Asia (20) and Australia (21). In the UK, ASQ^®^-3 is used in many local areas alongside professional judgement to decide which individual children are referred for extra support, in addition to collecting population level data (22).

In UK, there is a publicly available local-authority level dataset on health visiting, Health Visitor Service Delivery Metrics (HVSDM) which is submitted to the Office for Health Improvement and Disparities by local authorities (23-25). This data reports high coverage of ASQ^®^-3 e.g. in 2018/19, 71.4% of children aged 2½ years in the correct period had ASQ^®^-3 completed and ASQ^®^-3 was used in 90.4% of all 2-2½ year health and development reviews (figures for 2022/23 are 73.6% and 92.5% respectively (26). However, the Service Delivery Metrics cannot be used to analyse disparities as there is no individual or neighbourhood level data on ethnicity or deprivation and it cannot be linked to other administrative data.

Alternatively, we have an anonymised child-level data available in the CSDS which is supposed to hold complete data on health visiting, including ASQ^®^-3 data for every child in England who has had a 2-2½ year review. However, the CSDS has high levels of missingness which is due to the fact that providers remain at different stages of maturity in submitting their data to CSDS, including ASQ^®^-3 data (27). In previous work we found that, within 33 included local authorities, only 20-30% of children aged 2-3 years old in 2018/19 had a record of ASQ^®^-3 in CSDS (3). Thus, in this study, we identify a subset of CSDS ASQ^®^-3 data (between April 2018 and March 2021 (three financial years)) that is sufficiently complete to carry out research. We then use this subset to investigate child development at age 2-2^1/2^ years in England.

## Method

### Data source: Community Service Dataset (CSDS)

We used individual level ASQ^®^-3 data and demographic characteristics of the children captured in CSDS (28, 29) for the three financial years between April 2018 and March 2021. The ASQ^®^-3 data is entered into each local data systems by providers of health visiting (health visitors or the members of the health visiting team) and then uploaded monthly to the Community Services Dataset (CSDS) by local authority or NHS based data team with other data on community services, where it is collated at a national level (29).

We used SNOMED (Systematized Nomenclature of Medicine Clinical Terms) codes (30) to extract 2-2½ year child development outcomes done using 24, 27 or 30 month ASQ^®^-3 questionnaires (to cover all questionnaires used between the ages of 2-2½ years (14). We excluded duplicates meaning more than one ASQ-3 records per child by choosing the latest record. We also excluded records without demographic information in CSDS. More detailed process of identifying the eligible ASQ^®^-3 records is described in Appendix Figure 1.

We derived Lower Layer Super Output Area (LSOA) quintiles of deprivation from the Index of Multiple Deprivation (IMD) based on the child’s LSOA code. LSOA is a geographic hierarchy which is designed to improve the reporting of small area statistics in England and Wales and generally includes 400 to 1200 households or 1000 to 3000 people (31, 32).

### Outcome variable: ASQ^®^-3

#### Background

ASQ^®^-3 was developed to screen for developmental delay comprising 21 age specific questionnaires for children aged between 1 month and 66 months (5 ½ years) (33). Each questionnaire had 30 questions about the child’s development which were categorised into five domains with three response options (yes/sometimes/not yet). We used 2-2½ year child development outcomes done using 24-, 27- or 30-month ASQ^®^-3 questionnaires.

#### Domains

ASQ^®^-3 covers five key domains of developmental status of a child’s following areas (14, 23-25) (33):

- Communication - babbling, vocalizing, listening, and understanding
- Gross Motor: arm, body, and leg movements
- Fine Motor: hand and finger movements
- Problem Solving: learning and playing with toys
- Personal-Social: solitary social play and play with toys and other children.

Example questions of each of these domains can be found in Appendix Table 1.

#### Scores/Cut-offs

The score of each domain ranges between 0 and 60 which makes the total score 300 but the total score is rarely used since each domain has different cut-offs (14). For example, cut-off score for 27-month fine motor domain is 18.42 whereas it is 28.01 for Gross Motor domain. Different cut-offs for each ASQ^®^-3 domains are provided by the ASQ^®^-3 developers (14) that measure whether a child’s developmental status is at an expected level depending on the age. Cut-off score for each domain can be found in Appendix Table 2.

Based on the continuous ASQ^®^-3 scores in CSDS, we created a binary variable indicating whether a child reached expected or above level of development for all five domains (25). We used both binary cut-off variable and continuous ASQ^®^-3 score variable to understand child development at age 2-2^1/2^.

### Creating an analysis dataset with complete ASQ^®^-3 data

We assessed the completeness of ASQ^®^-3 data in CSDS for 2018/19-2020/21 at the local authority-quarter level by comparing the number of children with a completed ASQ^®^-3 to the number reported in the aggregate publicly available HVSDM. In previous work, we have found that HVSDM is accurate when compared to locally held data, which supports our use of HVSDM as reference data (3, 34). We had assumed that the quality of 2020/21 data might have been affected by the impact of the COVID-19 pandemic, but there were as many as valid ASQ^®^-3 records in 2020/21 compared to the previous two years. In our analysis dataset, we included those local authority-quarters where CSDS captured at least 85% the numbers of children who had an ASQ^®^-3 completed reported in the HVSDM. See Appendix Figure 2 for more information on methods for creating the analysis dataset which contained a subset of complete ASQ^®^-3 data in CSDS.

## Result

### Study sample: our ‘analysis dataset’

The analysis dataset included 293 local-authority-quarterly data points from 64 local authorities that had high agreement on ASQ^®^-3 data with the HVSDM between April 2018 and March 2021. The median number of quarters included in the analysis dataset was 4 (out of 12 quarters). There were some local authorities (n=9/64, 14.1%) that had just 1 complete quality quarter (i.e. which contribute only one quarter of data in our three-year study period (Appendix Figure 3). The local authorities in the analysis dataset were similar to all local authorities in England, based on region and urban/rural status but slightly less deprived (see Appendix Table 3).

Those children included in the analysis dataset (n=226,505) were slightly more deprived and less ethnically diverse than all children aged 2 years in England based on 2021 Census data published by the Office for National Statistics (ONS) (35) (Table 1). Our study sample contained a higher proportion of children in the most deprived IMD quintile (28.3%) than the national picture (24.8%). There was a higher proportion of children aged 2-2^1/2^ years with ‘white’ ethnicity in our analysis data set (78.9%) as in England as a whole, based on a comparison with 2021 census data (71.9%) (35).

**Table 1.** Characteristics of the children in the analysis dataset compared to all CSDS and ONS children and to all children with ASQ-3 records, % (2018/19 – 2020/21)

| Child characteristics | ONS | CSDS |  |  |
| --- | --- | --- | --- | --- |
|  | Aged 2-years (%)<br>(N=619,036) | All children aged 2-2<sup>1/2</sup> (%)<br>(N=2,264,995) | All children with ASQ®-3 (%)<br>(N= 432,910) | Children in the analysis dataset (%)<br>(N=226,505) |
| **Ethnicity** |  |  |  |  |
| White | 71.9 | 70.7 | 77 | 78.9 |
| Asian | 12.3 | 11.3 | 8.9 | 8.0 |
| Mixed | 8.0 | 8.7 | 7.5 | 7.0 |
| Black | 5.2 | 4.9 | 7.1 | 2.7 |
| Other | 2.7 | 4.4 | 3.5 | 3.4 |
| **Index of multiple deprivation (IMD) quintiles (LSOA)** |  |  |  |  |
| Most deprived | 24.8 | 25.9 | 27.9 | 28.3 |
| 2 | 21.7 | 21.9 | 20.1 | 19.2 |
| 3 | 19.2 | 19.3 | 18.8 | 18.3 |
| 4 | 17.8 | 17.2 | 16.9 | 16.6 |
| Least deprived | 16.6 | 15.7 | 16.4 | 17.6 |

### Child development at age 2-21/2 in England (Analysis dataset)

Among the 226,505 children in our analysis dataset, 86.2% had a record of expected or above level of development at age 2-2^1/2^ years based on ASQ^®^-3. This was slightly higher than the 83.4% (average of 2018/19-2020/21) reported by Public Health England (PHE) in ‘Child development outcomes at 2-2^1/2^ years’ (23-25).

We found that that 82.6% of children living in the most deprived neighbourhoods reached expected or above level of development based on their ASQ^®^-3 records in CSDS, compared to 85.0-89.7% of children living in all other levels of deprivation (Table 2).

**Table 2.**
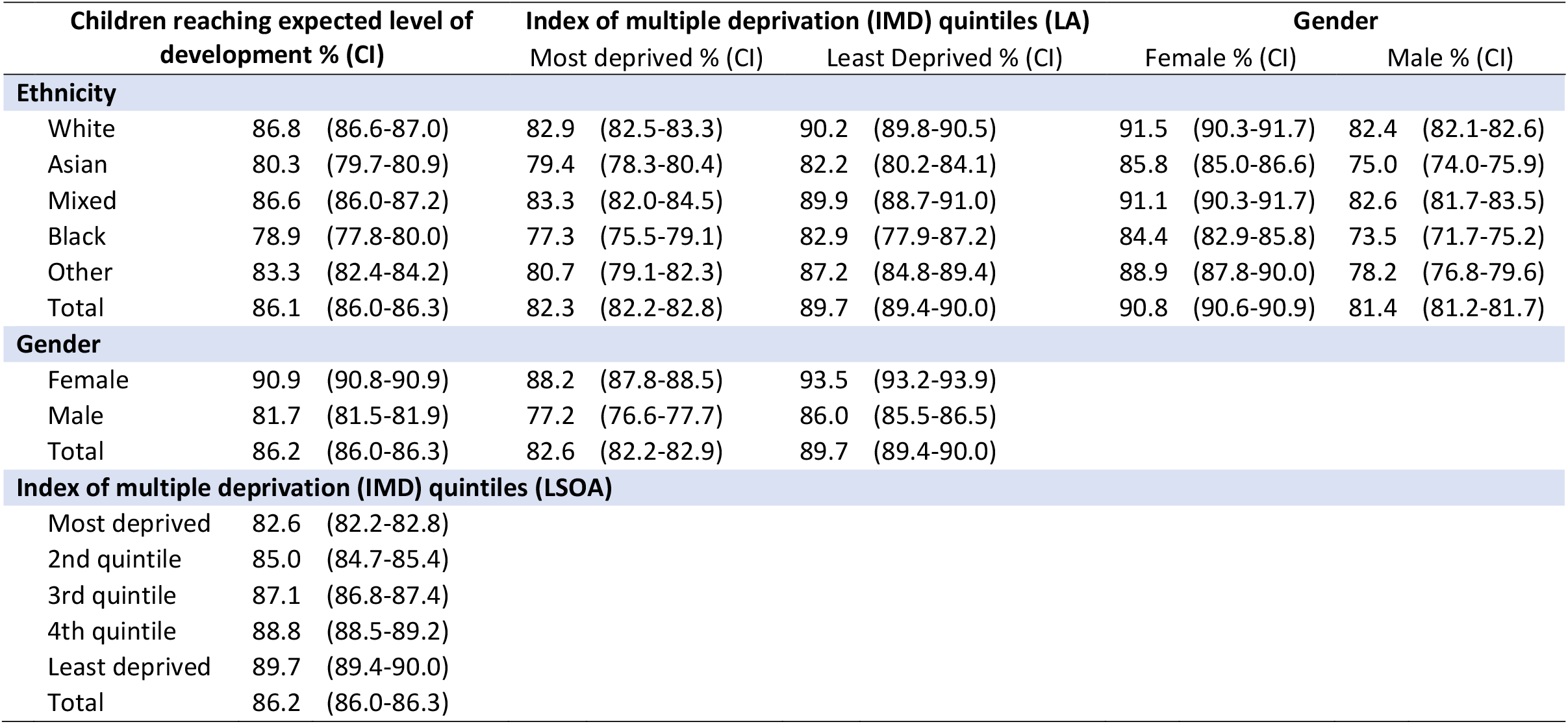
Variation in % children reaching at least expected level of development by characteristics (95% CI)

| Children reaching expected level of development % (CI) |  |  | Index of multiple deprivation (IMD) quintiles (LA) |  |  |  | Gender |  |  |  |
| --- | --- | --- | --- | --- | --- | --- | --- | --- | --- | --- |
|  |  |  | Most deprived % (CI) |  | Least Deprived % (CI) |  | Female % (CI) |  | Male % (CI) |  |
| Ethnicity |  |  |  |  |  |  |  |  |  |  |
| White | 86.8 | (86.6-87.0) | 82.9 | (82.5-83.3) | 90.2 | (89.8-90.5) | 91.5 | (90.3-91.7) | 82.4 | (82.1-82.6) |
| Asian | 80.3 | (79.7-80.9) | 79.4 | (78.3-80.4) | 82.2 | (80.2-84.1) | 85.8 | (85.0-86.6) | 75.0 | (74.0-75.9) |
| Mixed | 86.6 | (86.0-87.2) | 83.3 | (82.0-84.5) | 89.9 | (88.7-91.0) | 91.1 | (90.3-91.7) | 82.6 | (81.7-83.5) |
| Black | 78.9 | (77.8-80.0) | 77.3 | (75.5-79.1) | 82.9 | (77.9-87.2) | 84.4 | (82.9-85.8) | 73.5 | (71.7-75.2) |
| Other | 83.3 | (82.4-84.2) | 80.7 | (79.1-82.3) | 87.2 | (84.8-89.4) | 88.9 | (87.8-90.0) | 78.2 | (76.8-79.6) |
| Total | 86.1 | (86.0-86.3) | 82.3 | (82.2-82.8) | 89.7 | (89.4-90.0) | 90.8 | (90.6-90.9) | 81.4 | (81.2-81.7) |
| Gender |  |  |  |  |  |  |  |  |  |  |
| Female | 90.9 | (90.8-90.9) | 88.2 | (87.8-88.5) | 93.5 | (93.2-93.9) |  |  |  |  |
| Male | 81.7 | (81.5-81.9) | 77.2 | (76.6-77.7) | 86.0 | (85.5-86.5) |  |  |  |  |
| Total | 86.2 | (86.0-86.3) | 82.6 | (82.2-82.9) | 89.7 | (89.4-90.0) |  |  |  |  |
| Index of multiple deprivation (IMD) quintiles (LSOA) |  |  |  |  |  |  |  |  |  |  |
| Most deprived | 82.6 | (82.2-82.8) |  |  |  |  |  |  |  |  |
| 2nd quintile | 85.0 | (84.7-85.4) |  |  |  |  |  |  |  |  |
| 3rd quintile | 87.1 | (86.8-87.4) |  |  |  |  |  |  |  |  |
| 4th quintile | 88.8 | (88.5-89.2) |  |  |  |  |  |  |  |  |
| Least deprived | 89.7 | (89.4-90.0) |  |  |  |  |  |  |  |  |
| Total | 86.2 | (86.0-86.3) |  |  |  |  |  |  |  |  |

In our sample, a higher proportion of White (86.8%) and Mixed (86.6%) children reached an expected level of development compared to Black (78.9%) or Asian children (80.3%). Across all categories of recorded ethnicity, a high proportion of children living in the less deprived neighbourhoods reached an expected level of development than children of the same ethnicity living in the most deprived neighbourhoods (Table 4). A higher proportion of Girls (90.9%) reached expected level of development compared to boys (81.7%) regardless of ethnicity and deprivation (Table 2). Girls from the least deprived neighbourhoods (93.5%) were more likely to reach expected development than girls from the most deprived neighbourhoods (88.2%) and boys from both the least deprived (86.0%) and most deprived neighbourhoods (77.2%).

To find out whether this gender disparities on child development regardless of neighbourhood level deprivation shrunk depending on the local authority level deprivation, we categorised children into five local authority level IMD quintile groups (see Appendix Table 5). This tendency that girls from the most deprived neighbourhoods were more likely to reach expected level of development compared to the boys from the least deprived neighbourhoods was still found when children were grouped by their local authority level deprivation.

This gender disparities was also existed when we used ASQ^®^-3 score (ranges between 0-60) instead of a binary cut-off variable and split it into domains (Figure 1) and the differences also did not change by year (Appendix Figure 4). The gender gap was greatest for the communication and problem solving domains while it was less clear for fine motor domain.

**Figure 1.**
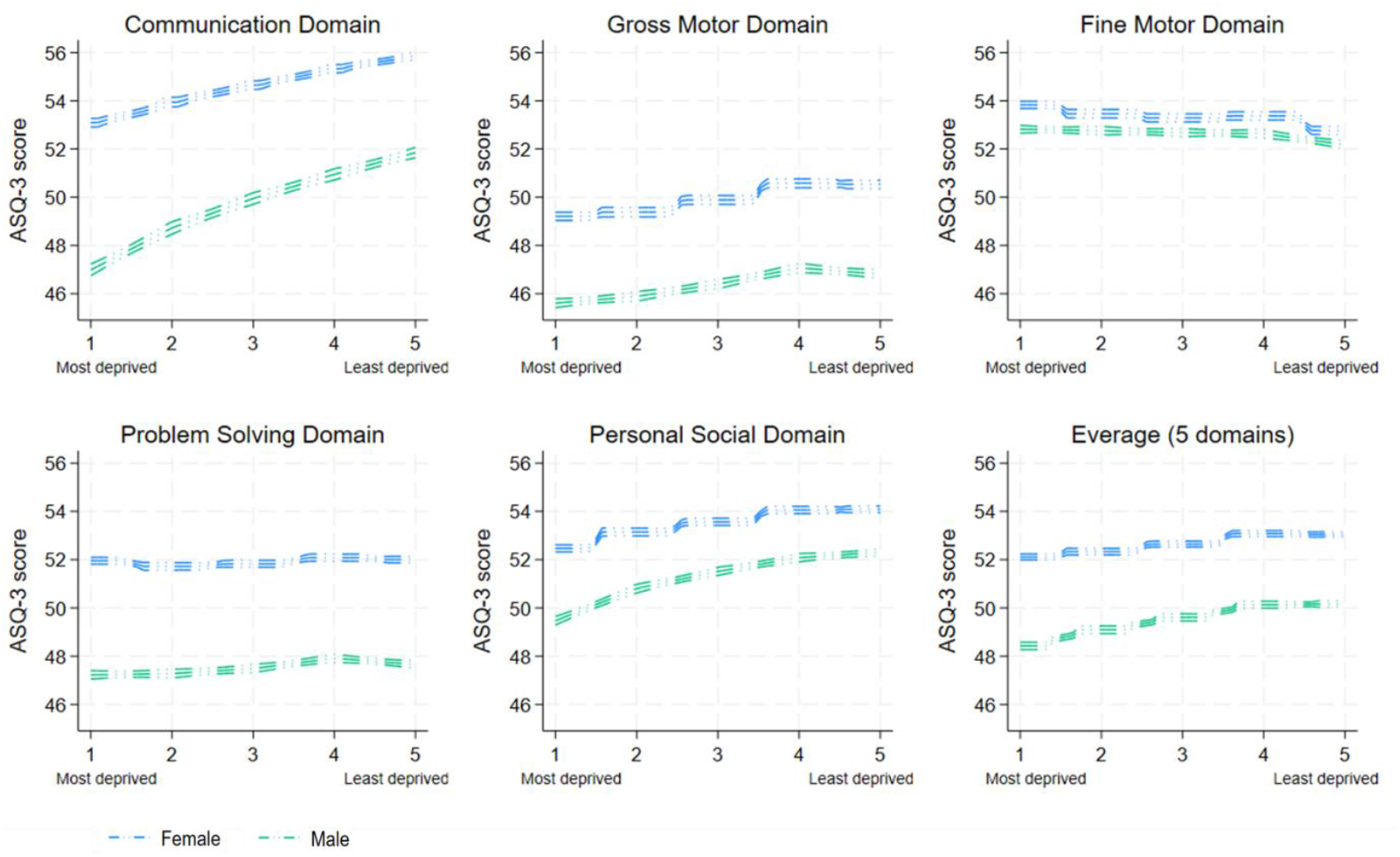
Average 27-month ASQ^®^-3 score by IMD (LSOA quintiles) and gender of the Child by domain Note: The two upper and bottle lines surrounding each gender lines are 95% CI.

Local authority level deprivation gradient in child development was also evident when looking at association between IMD and ASQ^®^-3 (Appendix Figure 5) although it did not seem to be as clear as it was found from the individual level analysis. When looking at the association between local authority level deprivation and child development (see Appendix Figure 6), the 3rd, 4th and least deprived local authority groups were similar in terms of average, but more variation was found in 3rd and 4th quintiles than least deprived local authorities. The most deprived quintile had a slightly higher average compared to the 2^nd^ quintile but there were more variations in child development in the 2^nd^ quintile.

## Discussion

### Main findings of this study

By assessing the quality of CSDS ASQ^®^-3 data, we derived an analysis dataset of 293 quarters of data from 64 local authorities, containing records for 226,505 children which we used to analyse which children in our sample reached expected levels of development at age 2-2½.

We found that in the most deprived neighbourhoods over 17% of children aged 2 years did not meet expected levels of development between April 2018 and March 2021, based on their ASQ^®^-3 records in CSDS. In our sample, almost a quarter (22.8%) of boys living in the most deprived neighbourhoods did not meet expected levels of development during this period. For local authorities with a high concentration of deprived neighbourhoods, this will represent a high number of young children not meeting expected development each year. Intervention and referral thresholds and pathways following ASQ^®^-3 are not clear – and we do not have a good idea of which of these children with lower than expected ASQ^®^-3 score received in terms of support and intervention and what that support or intervention looks like (36).

We found that there was a deprivation gradient, such that children from the most deprived neighbourhoods were the least likely to reach expected level of development. This result supports the findings of previous studies which revealed the deprivation gradient in child development (21, 37-39). For example, this is consistent with a finding of a study on local geographic variations in children’s school readiness which conducted multilevel analysis using the Department for Education Early Years Foundation Stage Profile (EYFSP) dataset measuring for 17 Early Learning Goals (ELGs) for 653,693 children aged between 4 and 5 years. This study found that children living in income deprived areas were the group with the lowest rates of being School Ready compared to the children from the less income deprived areas (37).

Regarding ethnicity, we found that white or mixed children were more likely to reach expected level of development compared to children with other ethnic background. This finding is in line with previous studies that revealed developmental gaps between children with different ethnic background (1, 37). A recent study on early child development which used the Millennium Cohort Study (MCS) also found that in the UK, white children (aged 3) were more likely to achieve higher development scores measured by the Bracken School Readiness test (40) and British Ability Scales II (41, 42) compared to those with other ethnic background (1). We found a striking gender gap in child development as girls were more likely to be assessed as meeting expected level of development compared to boys regardless of ethnicity or deprivation level. It is well documented that gender is strongly associated with early child development (16, 43, 44) which was found in many previous studies that girls in early ages outperformed boys (1, 21, 45). This gender disparities were evident in the studies that used the ASQ^®^-3 (17, 19, 21), the same tool as we used for our study, and also in other studies that used different tools, for example, The Caregiver-Reported Early Development Instruments (Credi) (38), Early Development Instrument (EDI) (44), The Education Early Years Foundation Stage Profile (EYFSP) (37) or Early Childhood Development Index (ECDI) (39, 46).

For example, McCoy and colleagues (38) used Credi (47) to measure the development of 8,022 children (aged under 3) from 17 low-, middle-, and high-income countries found that the score of girls was on average 0.08 SDs higher than boys. Strong gender effects on child development were also found by Purdam and colleagues (37) who used EYFSP dataset (48) based on 653,693 children (aged 4 and 5 years) in England. They found that girls from the most income deprived areas (decile 10) still showed better development than the boys living in less deprived areas (decile 4 to 9).

Studies that used ASQ^®^-3 also found significant gender effects on child development (19, 21)). For example, Veldman and colleagues (21) addressed the risk factors of child development using a sample of 701 pre-schoolers (3 to 5 years) living in low-income and remote communities in Australia and found that being a boy (odds ratio= 1.78) was one of the factors that was associated with a higher odds of children being delayed or at risk of gross motor delay.

### Implications

We found that there were 13.8% of the children who experienced development delays between April 2018 and March 2021. Even in the least deprived neighbourhoods with the higher % of children reaching expected development - e.g. 89.7% - there is still more than 10% of children not reaching this level. This highlights the importance of the appropriate support depending on the severity of the developmental delay.

Although CSDS ASQ^®^-3 is the only national level data that we can use to inform national level child development in England, this tool was not properly validated in the UK context (36) which in turn means that it is hard to know how to interpret the developmental differences between groups of children that we have found. For example, it was especially difficult to interpret a huge gender gap we found as we were cautious to conclude that the gender gap we found actually reflected the reality since it was not only unvalidated in the UK context but also not tested for whether any items on the ASQ^®^-3 exhibited gender bias. For example, Halpin and colleagues (49) validated the Early Childhood Development Index 2030 (ECDI2030) for children aged 24-59 months in Mexican- and Palestine-context where they also tested gender-based differential item functioning (DIF) to make sure the items on the ECDI2030 did not exhibit gender bias.

We also found it difficult to interpret the result using the cut-off for expected development. It is hard to say what the % of children not meeting expected development means given we have not had ASQ validated in the UK context. Thus, in our analysis, we have also used ASQ^®^-3 score to make sure the big gender gap we have found using a cut-off still existed when we used ASQ^®^-3 score.

In addition, ASQ^®^-3 is known to be sensitive in capturing severe forms of delay but tends to miss capturing minor forms of delay (36) which led us to think that there might be more children with minor developmental delay who still need support but were not captured by ASQ^®^-3.

### Strength and limitations

This study is the first study to evaluate the completeness of ASQ^®^-3 in the national administrative data in England (CSDS) and came up with the ASQ^®^-3 analysis dataset. This is important because researchers who are interested in using ASQ-^®^-3 data in CSDS can adopt ASQ^®^-3 analysis dataset which is a moderate size sample of local authorities/children in England. We also provided a baseline for the proportion of children in England reaching expected level of development between April 2018 and March 2021, across different levels of deprivation, ethnicities and in boys and girls using existing, routinely collected data.

Some of the limitations of this study stems from the incompleteness of the CSDS ASQ^®^-3 data itself. Although in theory all 2-2½ year olds in England should receive 2-year health visiting review using the ASQ^®^-3 questionnaire and the population measure of ASQ^®^-3 data should be available from the CSDS dataset, in reality ASQ^®^-3 data in CSDS is highly incomplete. Since our analysis dataset only included 64 local authorities with complete quality, we cannot generalise child outcome result to those local authorities that are not included in the analysis dataset. In the future, when the quality/completeness of the data is improved we will be able to have a clearer idea on child development at population level. Secondly, there were limited available child demographic information in CSDS which hindered an in-depth understanding of child development. Although we come up with some interesting findings on child development (e.g. strong gender gaps on child development) but it was difficult to address the relationship in depth as there were limited number of valid demographic factors in CSDS to further address this relationship. For example, in Cattan et al.’s (2023) study, the effect of ethnicity on child development shrunk for some groups (e.g. black-white gap in cognitive development) but did not change for other groups when other covariates (e.g. socio-economic and home learning environment) were included in the model. Since the important covariates that were known to affect child development (e.g. parental education level) were not included in our analysis, the result should be understood with caution.

We only had national level child development outcome measured when children were at age two which makes it difficult to address the trajectory of child development. As Peyre et al.’s (2019) study on sex differences in child development during preschool period in France found gender gap on child development shrunk as children got older but we cannot address this relationship and other factors’ long-term effects on child development using the available individual level administrative data in England.

## Conclusion

Early child development is critical as it shapes biological and psychological structures which in return affect their social and emotional wellbeing, cognitive skills and later educational achievement and long term academic success (2, 17). In order to allow government analysts and researchers to track trends in child development nationally (including variation over time and across groups and local areas), quantify need for preschool services and act as an outcome measure for evaluations of interventions in the early years, we need a population measure of early child development (19). This measure has been collected in England since 2015 in the form of universally administered ASQ^®^-3 at age 2-2^1/2^. However, to-date there have not been any analyses of this data across England looking at disparities between groups of children. Thus, we tested the quality of the individual level CSDS ASQ^®^-3 data which was highly incomplete and created the research ready quality analysis dataset (including 226,505 children from 64 local authorities). Using the analysis dataset, we addressed child development at age 2-2^1/2^ years and suggested that support is needed for at least 13.8% of the children who did not meet the expected level of development and especially for those who lived in the most deprived neighbourhoods and boys. We also found a huge gender gap regardless of ethnicity or deprivation, but the interpretation remained problematic as the lack of validation of the ASQ^®^-3 in the UK context.

We suggest that in order to fully understand child development in England, first ASQ^®^-3 data flow needs to be improved (50). Second, ASQ^®^-3 data needs to be standardised and validated in the UK context which is essential as the socio-demographic characteristics of the UK population are significantly different from that of the USA where the measure has been normed.

## Acknowledgements and funding

This work is supported by the National Institute for Health and Care Research (NIHR) Public Health Research Programme (NIHR129901) and Policy Research Programme (NIHR203450). The views expressed are those of the author(s) and not necessarily those of the NIHR or the Department of Health and Social Care. This research was supported in part by the NIHR Great Ormond Street Hospital Biomedical Research Centre. This research also benefits from and contributes to the NIHR Children and Families Policy Research Unit but was not commissioned through this Policy Research Unit (PR-PRU-1217-21301).

The views expressed are those of the author(s) and not necessarily those of the NIHR or the Department of Health and Social Care.

## Statement on Conflicts of Interest

The authors declare no conflicts of interest.

## Ethics Statement

This study has been approved by University College London Institute of Education (UCL IOE) Research Ethics Committee (1531).

## Data Availability Statement

Access to the CSDS was approved and provided by NHS England (NIC-393510 and NIC-381972). Health Visiting Service Delivery Metrics data are published by the Office for Health Improvement and Disparities and are openly available: data for 2018/19 (51) can be found at https://www.gov.uk/government/statistics/health-visitor-service-delivery-metrics-2018-to-2019, data for 2019/20 (52) can be found at https://www.gov.uk/government/statistics/health-visitor-service-delivery-metrics-experimental-statistics-2019-to-2020-annual-data and data for 2020/21 (53)) can be found at https://www.gov.uk/government/statistics/health-visitor-service-delivery-metrics-experimental-statistics-annual-data.

## Abbreviations

CSDS: Community Services Dataset
HVSDM: Health Visiting Service Delivery Metrics
IMD: The Index of Multiple Deprivation
LSOA: Lower Layer Super Output Area
ONS: Office for National Statistics

## Notes

### Competing Interest Statement

The authors have declared no competing interest.

### Funding Statement

This study was funded by the National Institute for Health and Care Research (NIHR) through the Children and Families Policy Research Unit (PR-PRU-1217-21301).
This work has also benefited from and contributed to National Institute for Health and Care Research (NIHR) Public Health Research Programme (NIHR129901) and Policy Research Programme (NIHR203450). This research was supported in part by the NIHR Great Ormond Street Hospital Biomedical Research Centre. The views expressed are those of the authors and not necessarily those of the NIHR or the Department of Health and Social Care.

### Author Declarations

University College London Institute of Education (UCL IOE) Research Ethics Committee (REC1725) gave ethical approval for this work.

