## Supplementary Material A for "Early child development in England: cross-sectional analysis of ASQ^®^-3 records from the 2-2^½^-year universal health visiting review using national administrative data (Community Service Dataset, CSDS)"

**Supplementary Material A. Tables and Figures**

Appendix Figure 1: Flow chart to identify the child development (ASQ&[reg]-3) records in Community Services Dataset from April 2018 to March 2021 for inclusion in our analysis dataset

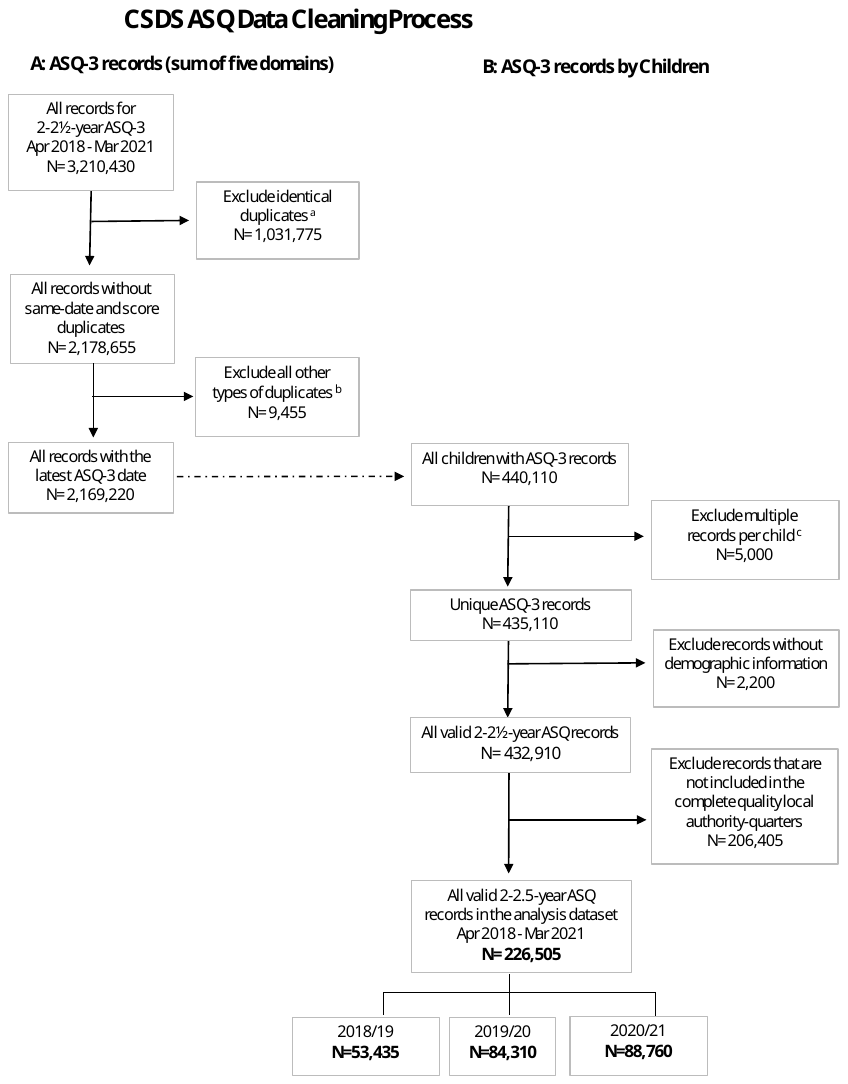

Notes: ^a^ 'identical duplicates' were records for which the ASQ&[reg]‐3 records per child were identical based on ASQ&[reg]‐3 domain, assessment date, and score. ^b^ Other types of duplicates include: (i) the same score recorded on different dates, (ii) different scores recorded on the same date, and (iii) different scores recorded on different dates. ^c^ Multiple ASQ&[reg]‐3 records per child in different years or done using different ASQ&[reg]‐3-months questionnaires.

Appendix Table 1 Example questions for each ASQ&[reg]-3 domain

| **Domain** | **Example question** |
| --- | --- |
| **Communication** | *‘Does your child use all of the words in a sentence (for example, “a,”*  *“the,” “am,” “is,” and “are”) to make complete sentences, such as “I*  *am going to the park,” or “Is there a toy to play with?” or “Are you*  *coming, too?”’* |
| **Gross motor** | *‘Does your child climb the rungs of a ladder of a playground slide and*  *slide down without help?’* |
| **Fine motor** | *‘Using child-safe scissors, does your child cut a paper in half on a more or less straight line, making the blades go up and down? (Carefully watch your child’s use of scissors for safety reasons.)’* |
| **Problem solving** | *‘When shown objects and asked, “What color is this?” does your child name five different colors, like red, blue, yellow, orange, black, white, or pink? (Mark “yes” only if your child answers the question correctly using five colors.)’* |
| **Personal-social** | *‘Does your child wash his hands using soap and water and dry off with a towel without help?’* |

Note: Example questions are copied from 48-month ASQ&[reg]-3 sample questionnaire [15] which is available from Ages and Stages Questionnaire webpage [59].

Appendix Table 2 Cut-off scores for each ASQ&[reg]-3 domain

| Domain | 24 month | 27 month | 30 month |
| --- | --- | --- | --- |
| Communication | 25.17 | 24.02 | 33.30 |
| Gross Motor | 38.07 | 28.01 | 36.14 |
| Fine Motor | 35.16 | 18.42 | 19.25 |
| Problem Solving | 29.78 | 27.62 | 27.08 |
| Personal-Social | 31.54 | 25.31 | 32.01 |

Source: Ages & stages questionnaires 3rd Edition (ASQ&[reg]-3) [15]

Appendix Figure 2 Completeness of CSDS ASQ&[reg]‐3 (2018/19 – 2020/21)

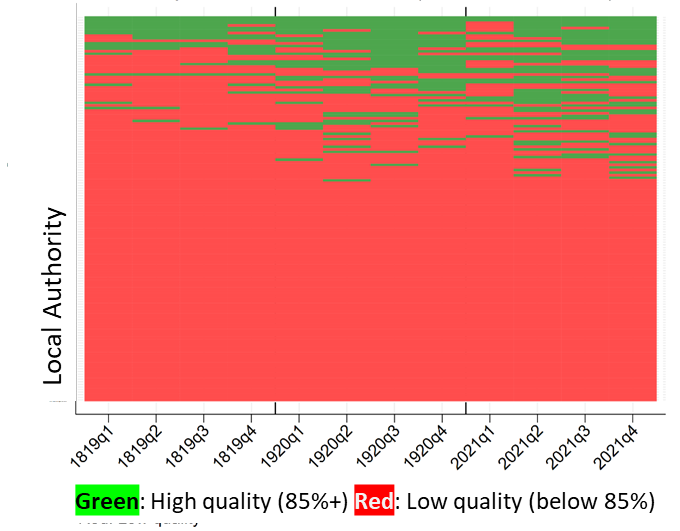

As there are 149 local authorities is England (health visiting in City of London is delivered by Hackney and in the Isles of Scilly it is delivered by Cornwall), we had reference data for 1,788 local authority-quarters (12 quarters x 149 local authorities).

In our previous studies, we have used a +/-15% margin to identify completed CSDS data for health visiting contacts [3, 30, 38, 60] based on the observed difference between counts of mandated contacts recorded in HVSDM and local health visiting activity data. However, in this study we assumed that if a child had plausible scores across all five domains of ASQ&[reg]-3 in the CSDS data that this was likely to accurately reflect that ASQ&[reg]-3 has been administered and we therefore included local authority quarters of data where ASQ&[reg]-3 records in CSDS exceeded numbers in the reference data (85%+). Where there were more ASQ&[reg]-3 records in CSDS than the HVSDM for a given local authority quarter, we included that local authority quarter of data (no upper limit).

Appendix Figure 3 Total N of quarters included in the analysis dataset (2018/19 - 2020/21)

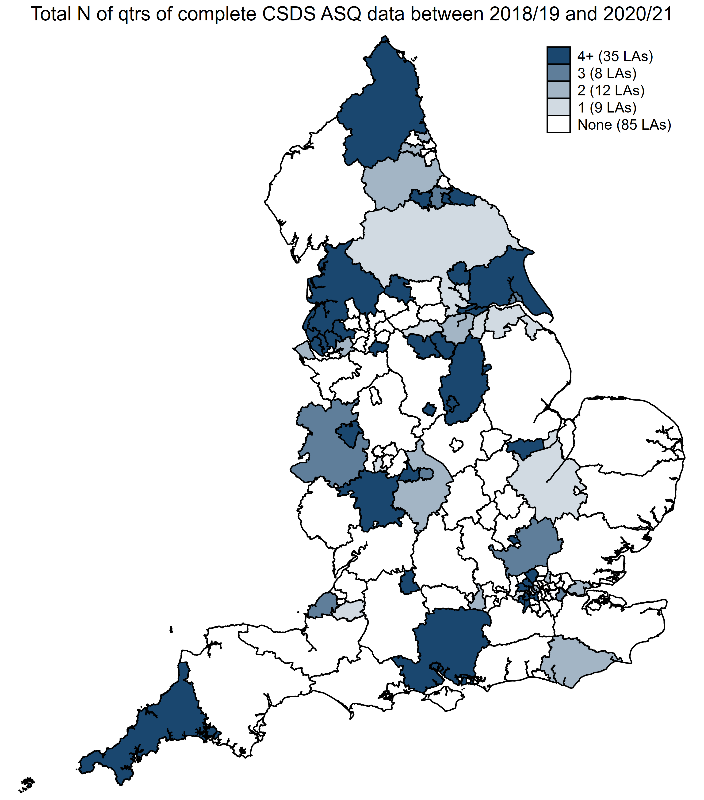

Appendix Table 3 Characteristics of local authorities included in the analysis dataset compared to all local authorities in England

| Local Authority Characteristics | | 149 local authorities in England  N (%) | | 64 local authorities included in the analysis dataset  N (%) | | P value comparing local authorities included with those not included in the analysis CSDS dataset |
| --- | --- | --- | --- | --- | --- | --- |
| **Region** | |  |  |  |  |  |
|  | East Midlands | 9 | (6.0) | 2 | (3.1) | 0.12 |
|  | East of England | 12 | (8.1) | 5 | (7.8) |  |
|  | London | 32 | (21.5) | 12 | (18.8) |  |
|  | North East | 12 | (8.1) | 8 | (12.5) |  |
|  | North West | 23 | (15.4) | 9 | (14.1) |  |
|  | South East | 18 | (12.1) | 5 | (7.8) |  |
|  | South West | 14 | (9.4) | 5 | (7.8) |  |
|  | West Midlands | 14 | (9.4) | 7 | (10.9) |  |
|  | Yorkshire and The Humber | 15 | (10.1) | 11 | (17.2) |  |
| **Geographical area classification ^a^** | |  |  |  |  |  |
|  | Predominantly Rural | 20 | (13.4) | 7 | (10.9) | 0.74 |
|  | Predominantly Urban | 108 | (72.5) | 48 | (75.0) |  |
|  | Urban with significant rural | 21 | (14.1) | 9 | (14.1) |  |
| **Income deprivation affecting children index (IDACI) quintiles ^b^** | | | | | | |
|  | Most deprived | 29 | (19.5) | 7 | (10.9) | 0.02 |
|  | 2 | 30 | (20.1) | 18 | (28.1) |  |
|  | 3 | 30 | (20.1) | 17 | (26.6) |  |
|  | 4 | 30 | (20.1) | 10 | (15.6) |  |
|  | Least deprived | 30 | (20.1) | 12 | (18.8) |  |
| **Index of multiple deprivation (IMD) quintiles ^b^** | | | |  |  |  |
|  | Most deprived | 29 | (19.5) | 7 | (10.9) | 0.05 |
|  | 2 | 30 | (20.1) | 17 | (26.6) |  |
|  | 3 | 30 | (20.1) | 17 | (26.6) |  |
|  | 4 | 30 | (20.1) | 12 | (18.8) |  |
|  | Least deprived | 30 | (20.1) | 11 | (17.2) |  |

Note: There are a total of 149 local authorities in the analysis as City of London is combined with Hackney and Isles of Scilly is combined with Cornwall. ***^a^*** Geographical area type was categorised using the Office for National Statistics (ONS) Rural Urban Classification lookup table for local authority areas [61]. ***^b^*** IDACI and IMD quintiles were based on the English Indices of Deprivation 2019, as published by the Ministry of Housing, Communities and Local Government [33].

Appendix Table 4 Percentage of children reaching expected level of development by deprivation and ethnicity; % (95% Confidence interval [CI])

|  | **Most  deprived**  **% (CI)** | **2nd quintile % (CI)** | **3rd quintile % (CI)** | **4th quintile % (CI)** | **Least  deprived**  **% (CI)** | **Total % (CI)** |
| --- | --- | --- | --- | --- | --- | --- |
| **White** | 82.9 | 85.9 | 87.8 | 89.4 | 90.2 | 86.8 |
|  | (82.5-83.3) | (85.5-86.3) | (87.4-88.2) | (89.0-89.8) | (89.8-90.5) | (86.6-87.0) |
| **Asian** | 79.4 | 80.5 | 79.9 | 81.5 | 82.2 | 80.3 |
|  | (78.3-80.4) | (79.2-81.8) | (78.3-81.4) | (79.5-83.4) | (80.2-84.1) | (79.7-80.9) |
| **Mixed** | 83.3 | 84.6 | 87.3 | 89.1 | 89.9 | 86.6 |
|  | (82.0-84.5) | (83.1-86.0) | (86.0-88.6) | (87.8-90.4) | (88.7-91.0) | (86.0-87.2) |
| **Black** | 77.3 | 79.3 | 81.1 | 79.3 | 82.9 | 78.9 |
|  | (75.5-79.1) | (77.0-81.5) | (78.3-83.7) | (75.3-82.8) | (77.9-87.2) | (77.8-80.0) |
| **Other** | 80.7 | 82.9 | 83.4 | 87.5 | 87.2 | 83.3 |
|  | (79.1-82.3) | (80.8-84.9) | (81.1-85.6) | (85.0-89.7) | (84.8-89.4) | (82.4-84.2) |
| **Total** | 82.5 | 85.0 | 87.1 | 88.8 | 89.7 | 86.1 |
|  | (82.2-82.8) | (84.7-85.4) | (86.8-87.4) | (88.5-89.2) | (89.4-90.0) | (86.0-86.3) |

Appendix Table 5 Percentage of children reaching expected level of development by local authority-level and LSOA-level Index of Multiple Deprivation (IMD) and gender; % (95% Confidence interval [CI])

| **Local authority level IMD** |  | **LSOA level IMD** | | | | | | | |
| --- | --- | --- | --- | --- | --- | --- | --- | --- | --- |
|  |  | **Most deprived quintile** | | | | **Least deprived quintile** | | | |
|  |  | Female % (CI) | | Male % (CI) | | Female % (CI) | | Male % (CI) | |
| **Most deprived** |  | 89.7 | (89.1-90.3) | 79.2 | (78.4-80.0) | 94.1 | (91.2-96.3) | 86.5 | (82.7-89.7) |
| **2nd quintile** |  | 86.7 | (86.0-87.3) | 74.4 | (73.6-75.2) | 93.4 | (92.3-94.3) | 84.4 | (82.9-85.8) |
| **3rd quintile** |  | 89.4 | (88.5-90.2) | 79.7 | (78.6-80.8) | 94.4 | (93.3-95.3) | 89.3 | (87.9-90.5) |
| **4th quintile** |  | 87.1 | (86.0-88.1) | 76.0 | (74.7-77.3) | 92.2 | (91.3-93.0) | 84.5 | (83.4-85.6) |
| **Least deprived** |  | 87.5 | (85.9-89.0) | 77.2 | (75.2-79.1) | 94.0 | (93.5-94.5) | 86.3 | (85.6-87.0) |

Appendix Figure 4 Average ASQ&[reg]-3 score by Index of Multiple Deprivation (quintiles) and gender of the child by ASQ&[reg]-3 domain and financial year

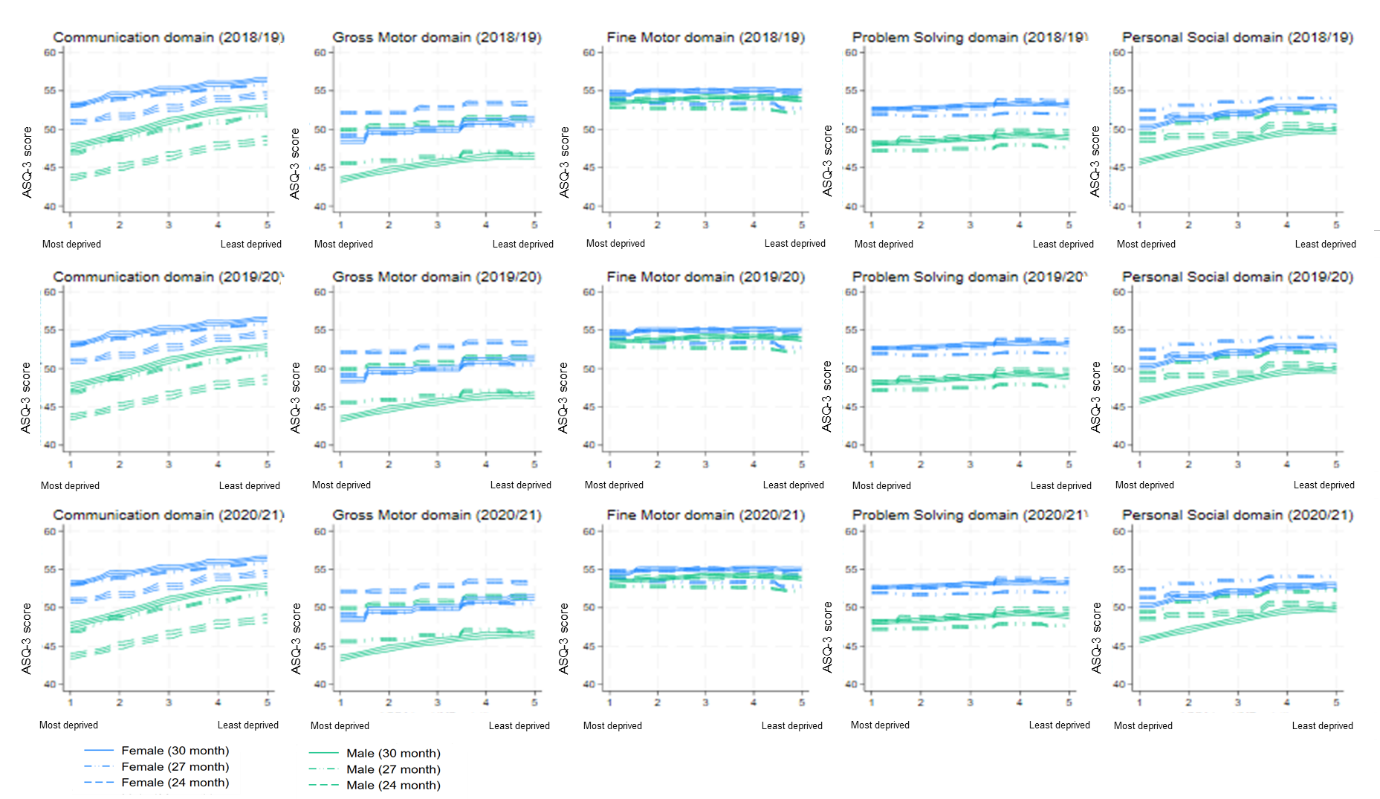

Appendix Figure 5 Association between local authority level deprivation (IMD score) and percentage of children meeting or exceeding the expected level of development (ASQ&[reg]-3)

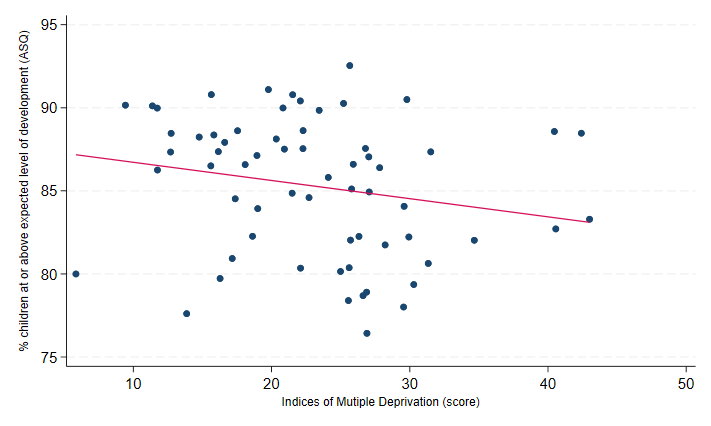

Appendix Figure 6 Association between local authority level deprivation (IMD quintile) and percentage of children meeting or exceeding the expected level of development (ASQ&[reg]-3)

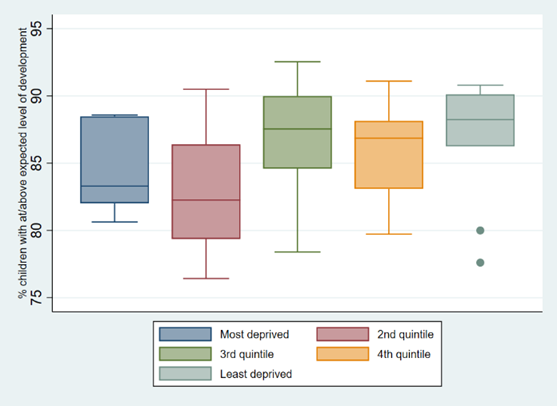
